# Arbidol treatment with reduced mortality of adult patients with COVID-19 in Wuhan, China: a retrospective cohort study

**DOI:** 10.1101/2020.04.11.20056523

**Authors:** Qibin Liu, Xuemin Fang, Lu Tian, Naveen Vankadari, Xianxiang Chen, Ke Wang, Dan Li, Xiyong Dai, Feng Xu, Lei Shen, Bing Wang, Li Yao, Peng Peng

## Abstract

**BACKGROUND:** The worldwide COVID-19 pandemic is increasing exponentially and demands an effective and promising therapy at most emergency.

**METHODS:** We have assembled a cohort consisting 504 hospitalized COVID-19 patients. Detailed information on patients’ characteristics and antiviral medication use during their stay at designated hospitals along with their pre and post treatment results were collected. The study objective is to evaluate the treatment efficacy of Arbidol, together with the concurrent drugs Oseltamivir and Lopinavir/Ritonavir on mortality and lesion absorption based on chest CT scan.

**FINDINGS:** The overall mortality rate was 15.67% in the cohort. The older age, lower SpO2 level, larger lesion, early admission date, and the presence of pre-existing conditions were associated with higher mortality. After adjusting for the patients age, sex, pre-existing condition, SpO2, lesion size, admission date, hospital, and concurrent antiviral drug use, Arbidol was found promising and associated with reduced mortality. The OR for Arbidol is 0·183 (95% CI, 0·075 to 0·446; P<0·001). Furthermore, Arbidol is also associated with faster lesion absorption after adjusting for patient’s characteristics and concurrent antiviral drug use (P=0·0203).

**INTERPRETATION:** The broad-spectrum antiviral drug Arbidol was found to be associated with faster

## Introduction

Since the first discovery of novel coronavirus disease 2019 (COVID-19) in Wuhan (China) in December 2019,^1^ more than 4.8 million people were infected and over 323,000 have died across the globe as of May 22, 2020,^2,3^ The world-wide rapid spread of the infection with limited therapeutic options is leading to a global health emergency. Meanwhile scientists and researchers are racing with time testing a huge variety of new and existing treatments. As of April 8, 2020, there were 388 ongoing studies on COVID-19 that are recorded in the U.S. National Library of Medicine of the NIH,^4^ including studies involving plasma, stem cells, and antiviral drugs, etc. For example, Liu et al (2020) presented preliminary results for 39 patients suggesting a potential benefit of convalescent plasma treatment in reducing mortality.^5^

Much attention has been drawn to a few promising candidates, namely Remdesivir, Lopinavir/Ritonavir, Hydroxychloroquine, Favipiravir and Umifenovir (Arbidol), however most completed studies so far had negative results. Mehra et al (2020) reported a multinational registry analysis of 96,032 patients and showed the use of Hydroxychloroquine or Chloroquine was associated with decreased in-hospital survival and increased ventricular arrhythmias;^6^ Geleris et al (2020) also reported the result of an observational study of 1446 patients showing Hydroxychloroquine was not associated with significant benefit on a composite endpoint of intubation or death;^7^ Cao et al (2020) reported that no benefit was observed with Lopinavir/Ritonavir treatment beyond standard care in a randomized, controlled, open-label trial with 199 patients.^8^ Chen et al (2020) reported a randomized clinical trial of 240 patients comparing Favipiravir with Arbidol, and found that Favipiravir did not significantly improve the clinical recovery rate at Day 7 as compared to Arbidol.^9^

Remdesivir has been regarded as one of the most promising drug candidates, with a total of three trial results becoming available so far.^10-12^ It was found to be superior to placebo in shortening the time to recovery, but its effect on the mortality was not statistically significant.^11^ We are still nowhere close to finding the best cure.

In the search for effective COVID-19 therapies, most researchers are targeting ACE2 and RdRp, but recent findings also suggest CD26 (DPP4) as another possible target.^13,14^ Viral entry is mediated via several pathways and it may not solely depend on the ACE2 which warrants testing other host targets. Recently, Vankadari (2020) reported the binding and mechanism of Arbidol action over the SARS-CoV-2 spike protein, which raises the significance of Arbidol being a possible drug candidate for COVID-19 infection.^15^

In this study, we aim to evaluate the clinical effect of Arbidol with a retrospective cohort of COVID-19 patients from Wuhan. We have also conducted molecular dynamics and time course simulation studies in order to understand the mechanism of the Arbidol drug action over the SARS-CoV-2 viral proteins.

## Methods

### Study design and participants

We have assembled a cohort consisting 504 in-patients with COVID-19 from three hospitals in Wuhan: Wuhan Pulmonary Hospital (WPH), Tongji Hospital, and Union Hospital. All three were designated COVID-19 hospitals during the outbreak. The study protocols were reviewed and approved by the Ethics committee of WPH (WPE 2020-12). Patient records and information were de-identified prior to the analysis.

Figure 1 shows the diagram of cohort design and patient selection. For WPH, we started with all COVID-19 patients admitted between December 13, 2019 and March 21, 2020 and excluded patients with ambiguous diagnosis as well as those with a primary cause of death unrelated to COVID-19. After further excluding 119 patients who remained being hospitalized by March 29, 2020, 373 patients from WPH were included. For Tongji Hospital, 100 COVID-19 patients admitted into two randomly selected hospital wards between February 1, 2020 and February 5, 2002 were included. Similarly, for Union Hospital, 31 COVID-19 patients admitted into a randomly selected ward between January 26, 2002 and February 24, 2002 were included. All patients in the study cohort have definitive outcomes, i.e., discharged or deceased. All COVID-19 infections are confirmed by Real-Time PCR (virus nucleic acid test).

**Figure 1:**
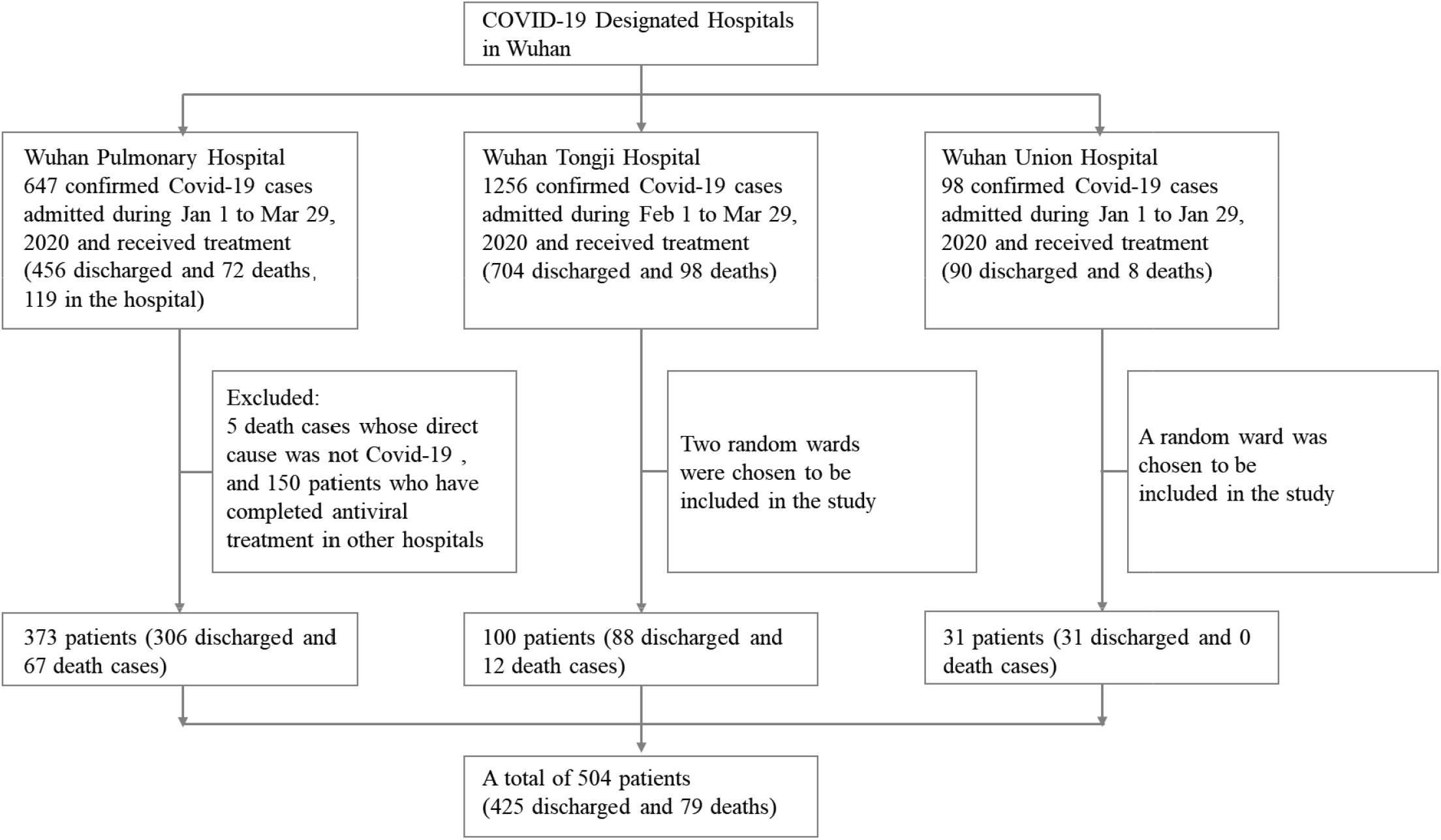
Diagram of cohort design and patient selection.

### Patients characteristics, medication and endpoints

We collected data on patients’ clinical outcomes (death or discharge) as well as basic characteristics, including age, sex, pre-existing conditions, SpO2 level at hospital admission, use of oxygen and ventilators during the hospital stay, and patient’s pre and post treatment lung CT scan images.

The drug treatment options used in all three hospitals followed the Chinese National Health Commission’s evolving guideline for the treatment of COVID-19, “New Corona Virus Pneumonia Diagnosis and Treatment Program”. The majority of the patients received Lopinavir/Ritonavir, Arbidol and Oseltamivir. With the development of the epidemic and the guideline updates, Hydroxychloroquine and Favipiravir were also used to a small number of patients. The prescription of antiviral medications and their administration time for each patient were extracted from electronic medical records with written consents from the clinicians. In this study, we specifically focus on three anti-viral medications, Arbidol, Oseltamivir, and Lopinavir/Ritonavir.

The primary endpoint is in-hospital death, with date of death extracted from the medical records. The secondary endpoint is the change in lesion size as measured by CT scans pre and post the antiviral treatment. CT scan images were read by certified thoracic radiologists at the respective hospitals to determine lesion size. The pre-treatment image was selected as the first CT image after the patient’s admission but before the initiation of the regular full-course antiviral treatment. The post-treatment image was selected as the last CT scan taken before patient’s death or discharge, which is normally after completing the antiviral therapy cycle. To ensure the comparability of the two CT images, we identified the plane with the largest lesion in the pre-treatment CT scan, and used the same plane in the post-treatment CT scan. The lesion in the selected plane was circled and size measured by the ImageJ software (Figure 2).^16^

**Figure 2:**
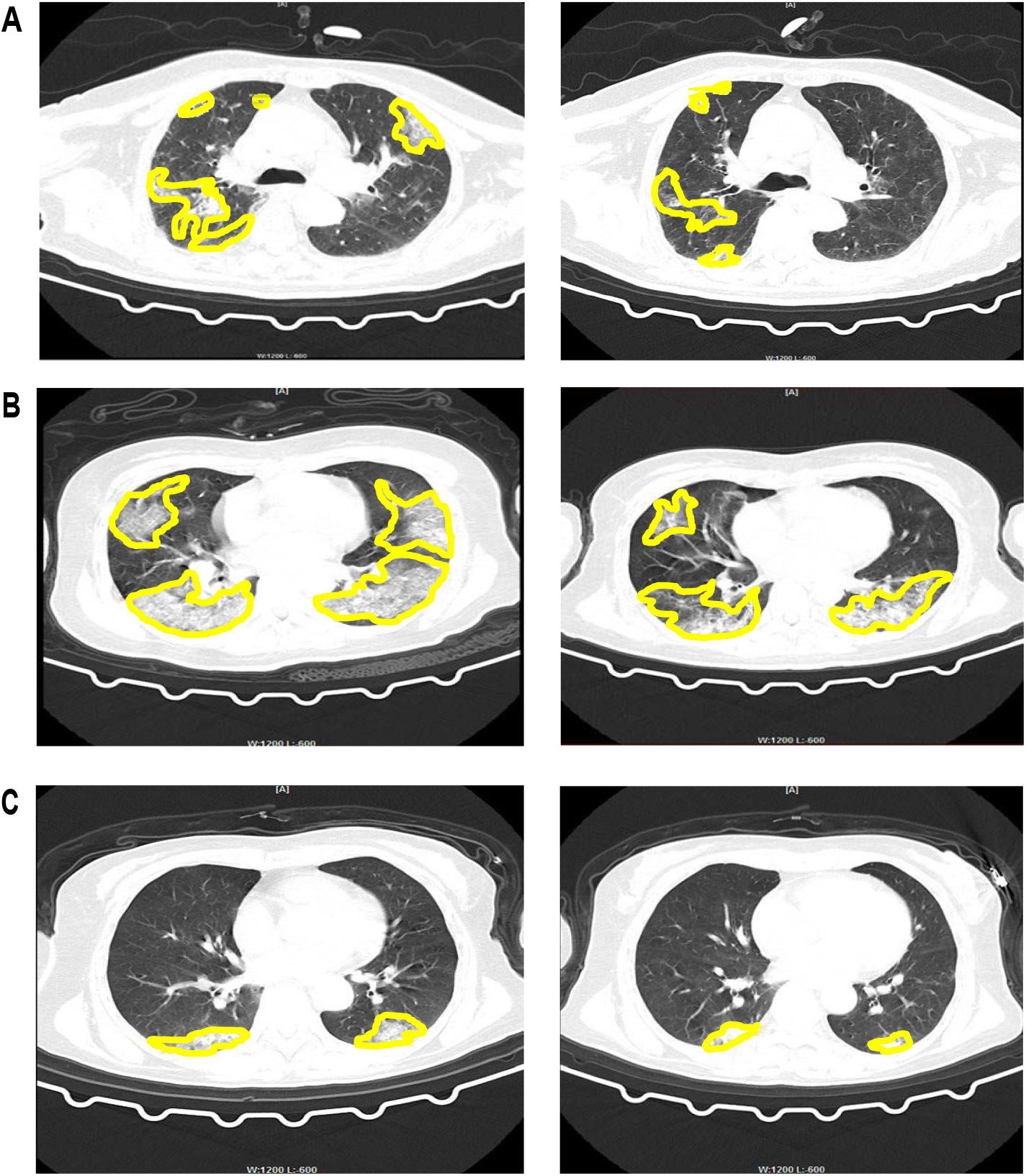
Chest CT scan images before and after the antiviral treatment. The shaded areas circled by the yellow line were the lesion areas. Using ImageJ, we were able to quantify the sizes of the lesions in terms of squared centimeters. **(A)** A male patient in his 50s with no underlying diseases who have received Arbidol alone. **(B)** A male patient in his 50s with no underlying diseases who have received Oseltamivir + Lopinavir/Ritonavir, since no patient has received Oseltamivir alone. **(C)** A male patient in his 50s with no underlying diseases who have received Lopinavir/Ritonavir alone.

### Statistical analysis

Continuous and dichotomous patient characteristics were summarized by their mean (standard deviation (SD)), and count (proportion), respectively. We compared the mortality rate between patients who received Arbidol, or Oseltamivir, or Lopinavir/Ritonavir and those who did not using Fisher’s exact test. We then employed the logistic regression to estimate the odds ratio (OR) associated with the treatment after adjusting sex, pre-existing condition, medication use, hospital, and log-transformed age, SpO2 level, admission date and the lesion size from the pre-treatment CT scan. Missing covariates were imputed by the medians of the observed values. We also repeated the logistic regression for patients in WPH only, where the potential sampling bias was the lowest. In addition, we performed Cox regression analyses with time dependent covariates, setting value to 0 and 1 before and after the medication prescription, respectively, to account for the timing of medication use.^17^ We also conducted linear regression analyses with the log-transformed ratio of lesion area absorption pre and post the antiviral treatments. A small value of one was added to all lesions sizes to avoid log-transformation of zero. The two-sided statistical significance level was set at 0.05. All statistical analyses were conducted using the R 3.3.1 software.

### Arbidol binding analysis

Docking studies with Arbidol and its binding analysis to SARS-CoV-2 spike glycoprotein was performed as described in Vankadari (2020).^15^ In brief, SwissDock (http://swissdock.ch/docking) server was used to dock the Arbidol into the published structure of SARS-CoV-2 spike glycoprotein trimer (PDB: 6VSB). Structural refinement was completed using Coot (www.mrc-imb.cam.uk/) to ensure no clashes in the side chain residues. Best dock score and highest binding free energies were taken into consideration to select the best-possible docking site and mode of interaction. Arbidol binding interface and the key residues interaction map was generated using the Maestro program. The molecular dynamics with time course simulation studies were performed using DynOmics server (www. http://gnm.csb.pitt.edu/). The final model with Arbidol docked in the homotrimer structure of SARS-CoV-2 spike protein and all other structural figures were visualized using PyMol.

### Role of the funding source

The authors report no funding.

## Results

### Cohort baseline characteristics

The cohort consists of 504 patients with confirmed COVID-19 infection from three hospitals: 373 patients from WPH, 100 patients from the Tongji Hospital, and 31 patients from the Union hospital. Table 1 summarizes main drug treatment options administrated during patient’s stay at the respective hospital. 257 patients received Arbidol (51·0%); 66 patients received Oseltamivir (13·1%); and 259 patients received Lopinavir/Ritonavir (51·4%). The majority of the patients received two or more different treatments during their hospitalization.

**Table 1.** Administered treatments for patients in three hospitals.

|  | **WPH**<br>**n=373** | **Tongji**<br>**n=100** | **Union**<br>**n=31** | **Total**<br>**n=504** |
| --- | --- | --- | --- | --- |
| Arbidol | 141 (37.8%) | 94 (94.0%) | 22 (71.0%) | 257 (51.0%) |
| Oseltamivir | 46 (12.3%) | 19 (19.0%) | 1 (3.2%) | 66 (13.1%) |
| Lopinavir/Ritonavir | 245 (65.7%) | 4 (4.0%) | 10 (32.3%) | 259 (51.4%) |
| Chloroquine | 4 (1.1%) | 0 (0.0%) | 0 (0.0%) | 4 (0.8%) |
| Hydroxychloroquine | 3 (0.8%) | 0 (0.0%) | 0 (0.0%) | 3 (0.6%) |
| Favipiravir | 4 (1.1%) | 0 (0.0%) | 0 (0.0%) | 4 (0.8%) |
| Ganciclovir | 28 (7.5%) | 0 (0.0%) | 2 (6.5%) | 30 (6.0%) |
| Glucocorticoid | 151 (40.5%) | 73 (73.0%) | 6 (19.4%) | 230 (45.6%) |
| Immunoglobulin | 117 (31.4%) | 25 (25.0%) | 10 (32.3%) | 152 (30.2%) |
| Albumin | 71 (19.0%) | 15 (15.0%) | 2 (6.5%) | 88 (17.5%) |
| Oxygen | 318 (85.3%) | 86 (86.0%) | 23 (74.2%) | 427 (84.7%) |
| Ventilation | 43 (11.5%) | 12 (12.0%) | 0 (0%) | 55 (10.9%) |

Among the 504 patients, 245 (48·6%) were female, and 262 (52·0%) had pre-existing conditions. The average age of the cohort was 59·5 (SD, 14·9) years old. The average Oxygen level (SpO2) at admission was 92·8% (9·3%). Patients who received Arbidol had slightly higher SpO2 level and smaller lesion area compared to other patients. Patients who received Oseltamivir were younger. In contrast, patients who received Lopinavir/Ritonavir had more pre-existing conditions. Patients admitted to the hospital early were more likely to receive Oseltamivir and Lopinavir/Ritonavir. Table 2 summarizes the baseline patients characteristics by hospital.

**Table 2.** Distributions of Baseline Characteristics by Medication and Hospital.

|  | WPH |  | Tongji |  | Union |  | Total |  |
| --- | --- | --- | --- | --- | --- | --- | --- | --- |
|  | Arbidol |  |  |  |  |  |  |  |
|  | Yes<br>n=141 | No<br>n=232 | Yes<br>n=94 | No<br>n=6 | Yes<br>n=22 | No<br>n=9 | Yes<br>n=257 | No<br>n=247 |
| Age<br>(year) | 61.3<br>(14.2)<sup>(1)</sup> | 60.3<br>(15.0) | 55.8<br>(14.6) | 68.5<br>(9.6) | 58.2<br>(17.3) | 46.4 (15.3) | 59.1<br>(14.8) | 60.0<br>(15.1) |
| Female | 72<br>(51.1%) | 112<br>(48.3%) | 41<br>(43.6%) | 3<br>(50.0%) | 12<br>(54.5%) | 5<br>(55.6%) | 125<br>(48.6%) | 120<br>(48.6%) |
| Pre-existing<br>conditions | 90<br>(63.8%) | 119<br>(51.3%) | 33<br>(35.1%) | 5<br>(83.3%) | 11<br>(50.0%) | 4<br>(44.4%) | 134<br>(52.1%) | 128<br>(51.8%) |
| SpO2 level<br>(%) | 93.6<br>(4.3) | 91.7<br>(12.2) | 93.9<br>(6.4) | 81.3<br>(14.9) | 95.8<br>(1.4) | 96.8<br>(1.1) | 93.9<br>(5.0) | 91.6<br>(12.1) |
| Lesion Size (cm<sup>2</sup>) | 56.1<br>(43.4) | 65.5<br>(48.4) | 46.8<br>(31.2) | 44.6<br>(19.7) | 55.4<br>(38.9) | 50.8<br>(42.6) | 52.7<br>(39.1) | 64.6<br>(47.9) |
| Admission<br>Date<sup>(2)</sup> (day) | 65.0<br>(13.4) | 61.2<br>(22.0) | 52.8<br>(1.4) | 52.5<br>(1.4) | 55.5<br>(7.3) | 55.4<br>(5.5) | 59.7<br>(11.7) | 60.8<br>(21.4) |
|  | Oseltamivir |  |  |  |  |  |  |  |
|  | Yes<br>n=46 | No<br>n=327 | Yes<br>n=19 | No<br>n=81 | Yes<br>n=1 | No<br>n=30 | Yes<br>n=66 | No<br>n=438 |
| Age<br>(year) | 59.1<br>(13.5) | 60.9<br>(14.8) | 52.1<br>(12.8) | 57.7<br>(14.9) | 66.0 | 54.4<br>(17.5) | 57.2<br>(13.6) | 59.9<br>(15.1) |
| Female | 24<br>(52.2%) | 160<br>(48.9%) | 9<br>(47.4%) | 35<br>(43.2%) | 1<br>(100%) | 16<br>(53.3%) | 34<br>(51.5%) | 211<br>(48.2%) |
| Pre-existing<br>Conditions | 27<br>(58.7%) | 182<br>(55.7%) | 4<br>(21.1%) | 34<br>(42.0%) | 1<br>(100%) | 14<br>(46.7%) | 32<br>(48.5%) | 230<br>(52.5%) |
| SpO2 level<br>(%) | 92.8<br>(6.4) | 92.4<br>(10.4) | 92.6<br>(7.7) | 93.3<br>(7.7) | 94.0 | 96.2<br>(1.3) | 92.8<br>(6.7) | 92.8<br>(9.6) |
| Lesion Size<br>(cm<sup>2</sup>) | 67.8<br>(46.1) | 60.3<br>(46.5) | 47.1<br>(39.5) | 46.7<br>(28.5) | 43.0 | 54.5<br>(39.8) | 61.5<br>(44.6) | 57.2<br>(43.4) |
| Admission<br>Date (day) | 42.2<br>(10.1) | 65.5<br>(18.5) | 53.2<br>(1.2) | 52.6<br>(1.4) | 63.0 | 55.2<br>(6.7) | 45.7<br>(10.0) | 62.4<br>(16.9) |
|  | Lopinavir/Ritonavir |  |  |  |  |  |  |  |
|  | Yes<br>n=245 | No<br>n=128 | Yes<br>n=4 | No<br>n=96 | Yes<br>n=10 | No<br>n=21 | Yes<br>n=259 | No<br>N=245 |
| Age<br>(year) | 60.3<br>(13.4) | 61.4<br>(16.9) | 39.2<br>(16.3) | 57.3<br>(14.2) | 53.5<br>(12.4) | 55.4<br>(19.5) | 59.7<br>(13.6) | 59.3<br>(16.2) |
| Female | 122<br>(49.8%) | 62<br>(48.4%) | 2<br>(50.0%) | 42<br>(43.8%) | 6<br>(60.0%) | 11<br>(52.4%) | 130<br>(50.2%) | 115<br>(46.9%) |
| Pre-existing<br>Conditions | 145<br>(59.2%) | 64<br>(50.0%) | 1<br>(25.0%) | 37<br>(38.5%) | 5<br>(50.0%) | 10<br>(47.6%) | 151<br>(58.3%) | 111<br>(45.3%) |
| SpO2 level<br>(%) | 92.5<br>(8.5) | 92.2<br>(12.4) | 95.0<br>(4.1) | 93.1<br>(7.8) | 96.9<br>(0.9) | 95.7<br>(1.4) | 92.7<br>(8.3) | 92.9<br>(10.2) |
| Lesion Size<br>(cm<sup>2</sup>) | 61.6<br>(47.2) | 61.0<br>(44.9) | 22.6<br>(17.3) | 48.0<br>(31.0) | 44.4<br>(30.8) | 59.0<br>(42.6) | 60.2<br>(46.6) | 55.1<br>(39.5) |
| Admission<br>Date (day) | 58.1<br>(15.4) | 71.4<br>(22.7) | 53.0<br>(1.6) | 52.7<br>(1.4) | 52.8<br>(5.0) | 56.7<br>(7.2) | 57.8<br>(15.0) | 62.8<br>(18.8) |

### The association between the treatments and mortality

For the cohort of 504 patients, the overall mortality rate was 15·67%. The mortality rate was 9·39% among females, 21·62% among males. The mortality rate was 20·23% among patients with pre-existing conditions, and 10·74% among patients without pre-existing conditions. The mortality rate was 17·96% in WPH, 12·00% in Tongji Hospital, and 0% in Union hospital. In this cohort, older age, lower SpO2 level at admission, larger lesion, and early admission date were associated with higher mortality.

The mortality rate was 7·00% among patients who received Arbidol compared to 24·70% among patients who did not. The odds ratio (OR) was 0·230 (95% confidence interval (CI), 0·124 to 0·411) favoring Arbidol. On the other hand, the mortality rate was 12·12% among patients who received Oseltamivir, compared to 16·21% among patients who did not. The OR was 0·713 (95% CI, 0·282 to 1·589) favoring Oseltamivir. The mortality rate was 14·29% among patients who received Lopinavir/Ritonavir, compared to 17·14% among patients who did not. The OR was 0·806 (95% CI, 0·483 to 1·341) favoring Lopinavir/Ritonavir. After adjusting for sex, pre-existing conditions, log(age), log(SpO2), log(lesion size), log(admission date) and hospital (model 1) using a multivariate logistic regression model, the OR was 0·169 (95% CI, 0·071 to 0·398) for Arbidol, 0·212 (95% CI, 0·072 to 0·623) for Oseltamivir, and 0·363 (95% CI, 0·165 to 0·795) for Lopinavir/Ritonavir. After further adjustment of concurrent antiviral medications (model 2), the beneficial effect of Arbidol and Oseltamivir have remained statistically significant: the adjusted OR was 0·183 (95% CI, 0·075 to 0·446; p<0·001) for Arbidol and 0·220 (95% CI, 0·069 to 0·707; P=0·011) for Oseltamivir (Figure 3A).

**Figure 3:**
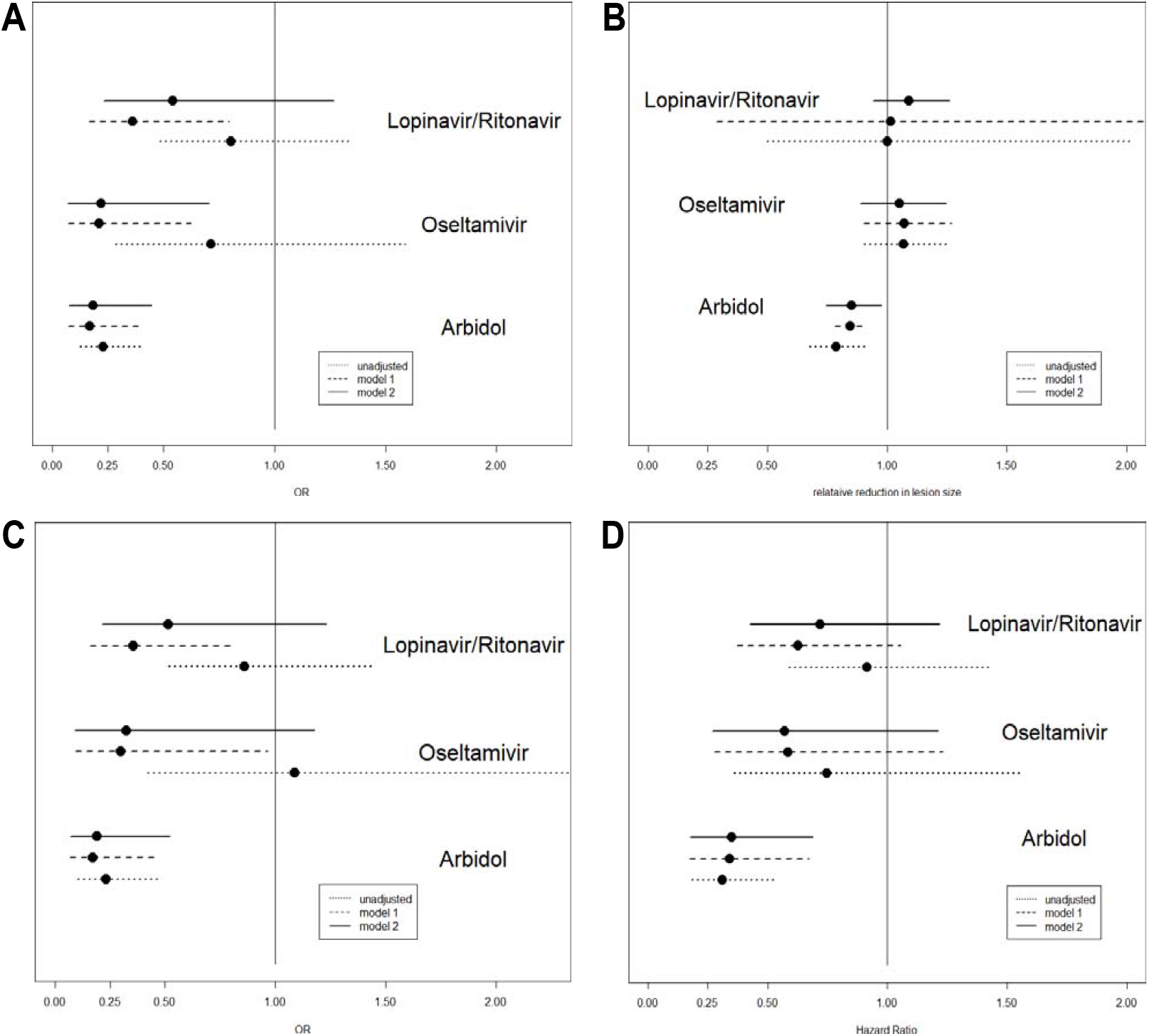
**(A)** The estimated OR and associated 95% CI for Arbidol, Oseltamivir and Lopinavir/Ritonavir. **(B)** The estimated effect on relative change in lesion size and associated 95% CI for Arbidol, Oseltamivir and Lopinavir/Ritonavir. An effect less than 1 suggests that more relative reduction in lesion size is associated with the medicine of interest. **(C)** The estimated OR and associated 95% CI for Arbidol, Oseltamivir and Lopinavir/Ritonavir among 373 patients from WPH. **(D)** The estimated hazard ratio and associated 95% CI for Arbidol, Oseltamivir and Lopinavir/Ritonavir based on the Cox regression analyses with time dependent covariates accounting for timings of medication use. Model 1: adjusting for sex, pre-existing condition, log(age), log(SpO2), hospital, log(lesion size) and log(admission date); Model 2: adjusting for confounders in Model 1 and medication use (Arbidol, Oseltamivir, and Lopinavir/Ritonavir).

### The association between the treatments and the reduction of lung lesion sizes

There were 326 survivors with two available CT scans. The average reductions in lesion size were 46·43% (SD: 29·00%) among the 209 patients who received Arbidol and 36·80% (SD: 24·95%) among the 117 patients who did not. The average reduction among the 55 patients who received Oseltamivir was less than that among the 271 patients who did not (41·18% vs 43·34%). The reduction among the 186 patients who received Lopinavir/Ritonavir was also less than the 140 patients who did not (37·26% vs. 50·56%). After adjusting for patients’ characteristics and concurrent antiviral drug use using a multivariate linear regression model, the ratio of the lesion size (post-treatment to pre-treatment) among patients who received Arbidol was 85·20% (95% CI, 74·47% to 97·48%; P=0·0203) of that among patients who did not, suggesting a faster lesion absorption. Figure 3B summarized the linear regression analysis results for all three antiviral drugs. Figure 4A shows the boxplots of the lesion absorptions in the stratified subgroups of patients by the three antiviral drugs, with patients who received Arbidol (dark turquoise) showing more absorption than patients who did not (light turquoise).

**Figure 4:**
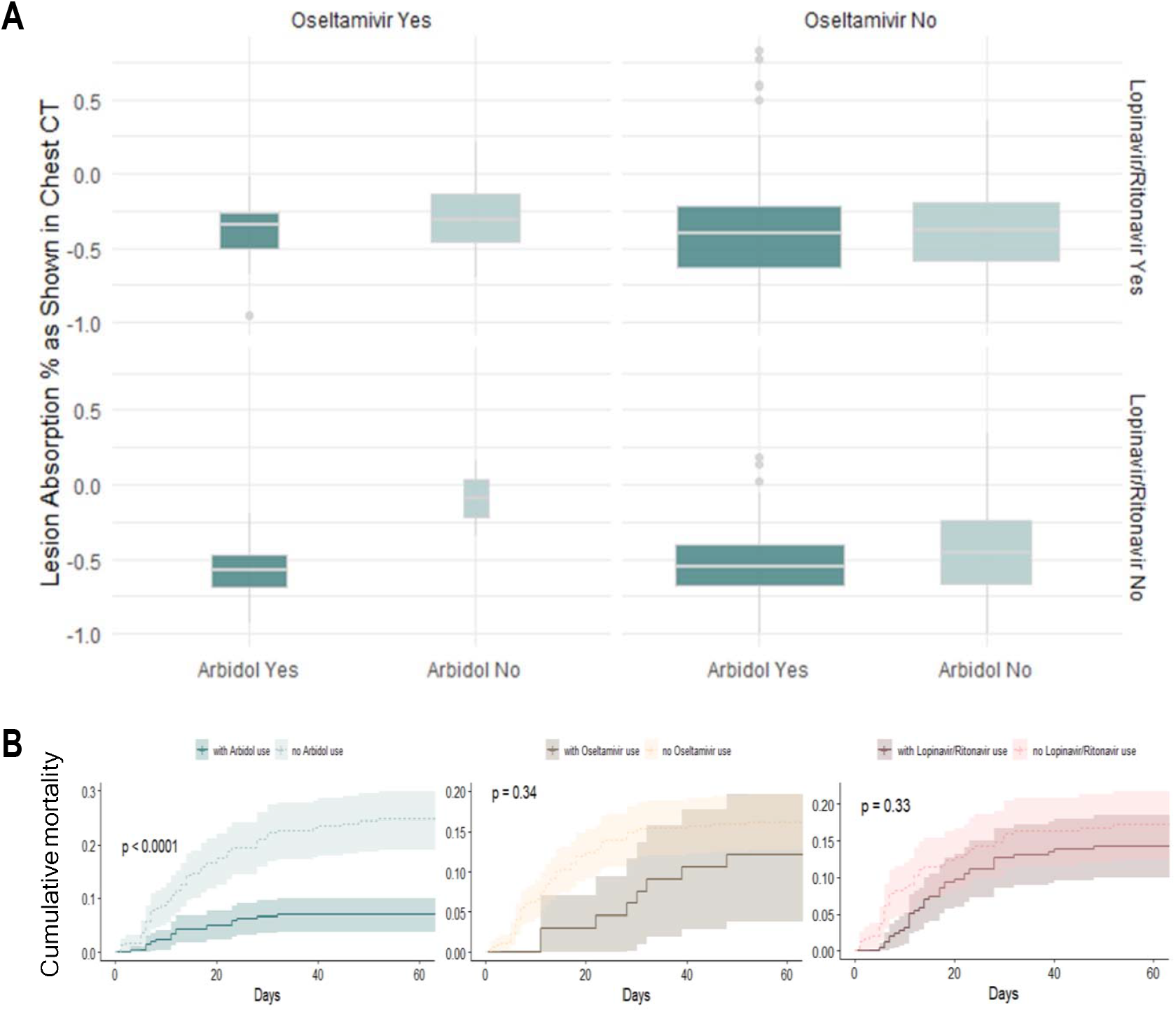
**(A)** Boxplot of the lesion absorption rate before and after the antiviral treatment, as shown in chest CT scan, compared among Arbidol, Oseltamivir, and Lopinavir/Ritonavir groups. In these boxplots we included CT lesion measures from all patients (deceased and discharged). **(B)** Cumulative mortality rate curves by medication use (Arbidol, Oseltamivir, and Lopinavir/Ritonavir). P-value of the log-rank tests are displayed and the shaded area shows the 95% confidence intervals

### Subgroup analysis in WPH

For the 373 patients in WPH, the mortality rate was 7·09% among patients who received Arbidol vs. 24·57% among patients who did not (OR=0·234, 95% CI, 0·103 to 0·481, P<0·001). After adjusting for sex, pre-existing condition, log(age), log(SpO2), log(lesion area), log(admission date), and concurrent antiviral drug use using a multivariate logistic regression model, Arbidol was associated with significantly reduced mortality (OR, 0·193; 95% CI, 0·071 to 0·520, P=0·001). The effect of Oseltamivir was marginally significant (OR, 0·326; 95% CI, 0·090 to 1·177; P=0·087). Figure 3C summarized detailed results.

### Survival analysis

Among the patients who received Arbidol, the median prescription time was 1·50 (IQR: 0·50-2·50, range, 0·50-31·50) days after admission. For Oseltamivir and Lopinavir/Ritonavir, the median prescription time was 0·50 (IQR: 0·50 to 1·50, range, 0·50-25·50) and 0·50 (IQR: 0·50 to 2·50, range, 0·50-32·50) days after admission, respectively. After adjusting for baseline characteristics and concurrent antiviral drug use using a multivariate cox proportional hazard ratio model, the hazard ratio was 0·350 (95% CI, 0·177 to 0·689; P=0·002) for Arbidol, 0·571 (95% CI, 0·269 to 1·211) for Oseltamivir, and 0·720 (95% CI, 0·426 to 1·218) for Lopinavir/Ritonavir based on Cox regression stratified by hospitals (Figure 3D). Figure 4B shows the cumulative mortality curves for the three antiviral drugs. There is a significant difference between the mortality curve for patients who received Arbidol versus patients who did not (P < 0·001) whereas the differences are not statistically significant for Oseltamivir (P = 0·34) and Lopinavir/Ritonavir (P = 0·33).

### Arbidol and SARS-CoV-2 spike glycoprotein interaction

It is evident from the previous studies that Arbidol is potentially effective against SARS-CoV-2.^13^ The recent structural studies of Arbidol in complex with SARS-CoV-2 spike glycoprotein underlines its role in impeding trimerization,^15^ which is essential for virulence. Here we address how this drug disrupts the viral spike glycoprotein through comprehensive drug interaction via molecular dynamics and time course simulation studies. As shown in Figure 5A, each monomer of spike glycoprotein consists of large surface for drug interaction and it could completely accommodate the drug in the trimer interface (Figure 5A and 5B). The time course simulation and drug interaction studies show that the Arbidol disrupt the interface in all directions and its binding affinity found to be very high (Figure 5C). The interaction is not only mediated by hydrogen bonding and van-der Wall forces but also by several polar residues bridge with the spike glycoproteins (Figure 5D). Furthermore, the dynamic simulation and domain movement or oscillation studies show that binding of Arbidol to the spike glycoprotein increases the B-factor (stability factor) (Figure 5E), suggesting protein is under higher movement and is less stable or tend to dissociate from other monomers. These structural and molecular dynamic studies further corroborate our clinical finding of CT scans showing greater viral clearance observed with Arbidol treatment.

**Figure 5:**
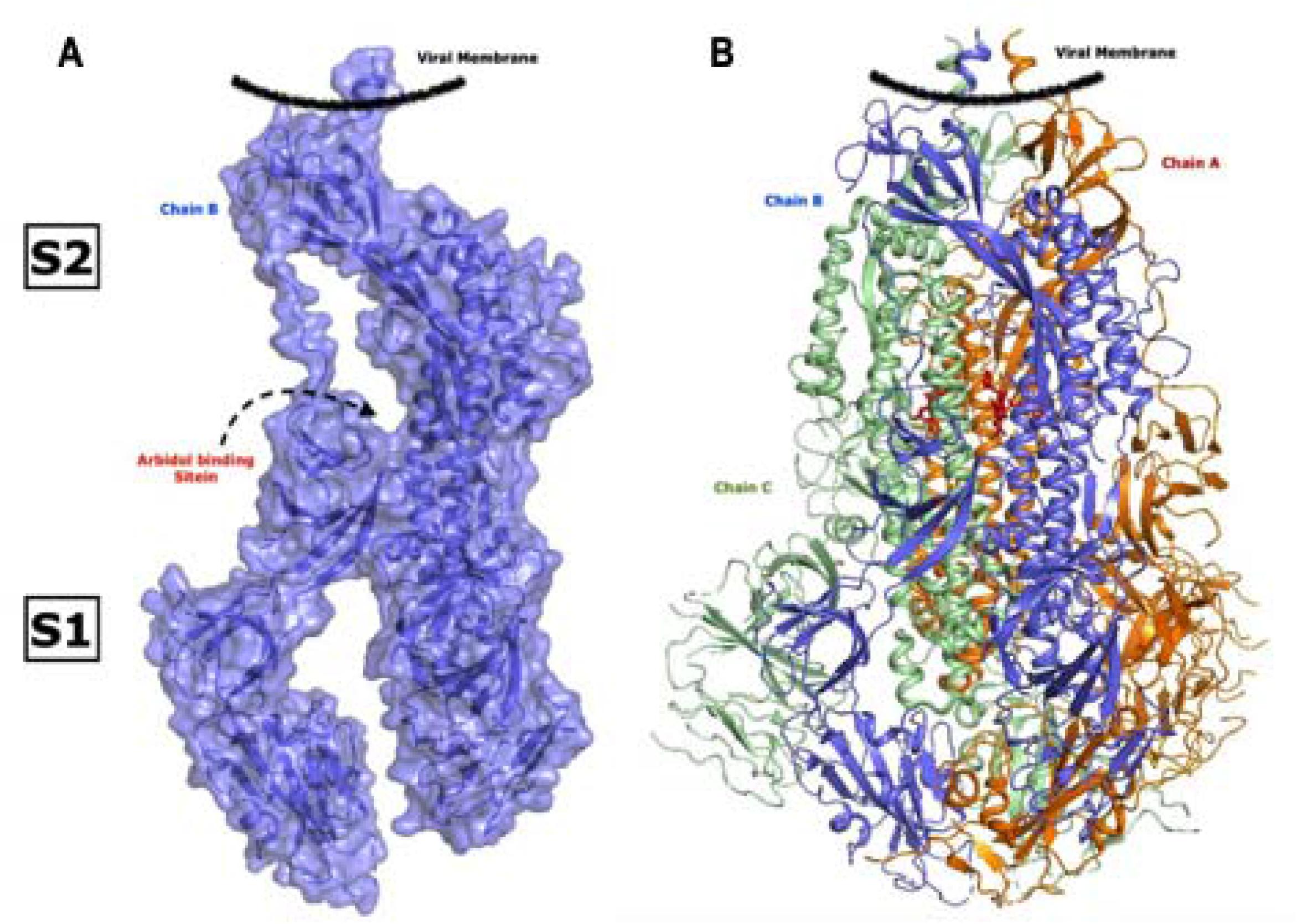
Surface and ribbon model showing the full length monomer **(A)** and homo-trimer **(B)** structure of SARS-CoV-2 spike glycoprotein. Position of S1 and S2 domains are labelled and bound Arbidol with high affinity at the center is shown in sticks. Each monomer of homo-trimer is shown in different colour.

## Discussion

The study showed promising treatment effect of Arbidol on mortality and lesion absorption. On the other hand, the benefit of Lopinavir/Ritonavir is inconclusive, which is consistent with the previous finding of Cao et al. (2020), where Lopinavir/Ritonavir has reduced the mortality from 25·0% to 19·2% in a randomized clinical trial, but failed to reach the statistical significance threshold.^8^ Recently, Xu et al, (2020) reported that the virologic conversion rate in 49 patients who received Arbidol was significantly higher than that in 62 patients who received the standard care (59·2% vs 40·3%, P=0·048).^19^ In this study, the patients who received Arbidol also had faster lung lesion absorption (55·1% vs 32·2%). Despite the limited sample size, their results further corroborated our findings. The dual role of Arbidol in inhibiting the fusion between the viral envelope and target host cell membrane as well as anti-inflammation could be responsible for its observed efficacy.^20, 21^

Oseltamivir is normally prescribed for the treatment of influenza and has no known effect on COVID-19 patients. However, some patients have been infected by both COVID-19 and Influenza,^22^ which may exacerbate their clinical conditions. It is very likely that a combination of drugs or therapy including Oseltamivir can help this subgroup of patients due to the effectiveness of Oseltamivir in treating severe illness caused by Influenza. In addition, SARS-CoV-2 and influenza viruses share a similar region of viral spike protein, which may also explain the potential effect of Oseltamivir in treating COVID-19 patients.^13^ There are some important limitations in the current study. First, the study is not a randomized clinical trial, and therefore the observed treatment benefit of Arbidol and Oseltamivir may be due to confounding effect. The analysis has adjusted for several known confounders including age, comorbidities, admission date, and disease severity measured through SpO2 and CT scan. Furthermore, given the size of the estimated treatment effect, the unmeasured confounding effect needs to be very strong to completely account for this benefit. Second, we didn’t account for the effect of important supporting treatment such as oxygen and ventilator use. Their availability and deployment may affect the estimated treatment effect. Third, although the cohort size is 504, the patients from Tongji and Union Hospitals are not necessarily the most representative samples of patients admitted into these hospitals. For example, ICU patients in these two hospitals are not included. We have also excluded surviving patients not yet discharged from WPH by March 29, 2020, and consequently, the observed mortality rate among patients from WPH is substantially higher than that from Tongji and Union Hospitals. These sampling biases may affect the interpretation and generalizability of our findings. For example, the reported mortality rate doesn’t represent the absolute mortality rate in the patients admitted into relevant hospital. Furthermore, there are only 33 patients who received both Arbidol and Oseltamivir in the entire cohort, which limits the reliability of the estimated treatment benefit associated with the combination therapy. On the other hand, despite the limited sample size and selective sampling, all identified risk factors in the current study are consistent with other studies,^22,23^ which indirectly supports our findings on the treatment efficacy. The association between admission date and mortality can also be explained by the availability of medical resources. Lastly, regarding the adverse events, nausea was observed in a small group of patients who received Arbidol. Loss of appetite was observed in patients who received Oseltamivir. Whereas Diarrhea, nausea, vomiting, loss of appetite and decreased sleep quality were observed in patients who received Lopinavir/Ritonavir. Due to the difficulty in accurate and complete documentation of all adverse events during the pandemic, the study team did not summarize these data in this report. Nevertheless, it is known that the severe adverse effects associated with the use of Arbidol and Oseltamivir have been very rare.^21,24^

Arbidol is originated from Russia as a broad-spectrum antiviral drug and is now widely used in Russia as an OTC drug and in China as a prescription drug. Although Arbidol was reported to effectively shorten the duration of influenza and prevent the development of post-influenza complications, reduce the re-infection risk based on multiple clinical trials conducted in Russia and China,^25^ due to lack of detailed information regarding these trials, WHO suggested that the results should be interpreted with caution.^26^ Consequently, Arbidol has not received FDA approval in the United States and the use of Arbidol in other parts of the world has been limited. However, while drafting this manuscript, we have found that eight clinical trials (NCT04350684, NCT04286503, NCT04260594, NCT04273763, NCT04323345, NCT04261907, NCT04306497, NCT04333589) in China, Iran and Egypt involving Arbidol in treating COVID-19 patients were registered at www.clinicaltrial.gov.

In conclusion, our clinical and molecular biological findings suggest that Arbidol might be a potentially effective treatment for COVID-19. Further clinical studies including RCTs are warranted for statistical and clinical evaluation of its efficacy.

## Data Availability

The data used to support the findings of this study are available from the corresponding author upon request.

## Notes

### Competing Interest Statement

The authors have declared no competing interest.

